# Employing Data-Driven Techniques to Explore the Lay Public’s Health Concerns with Vaping E-Cigarettes

**DOI:** 10.64898/2026.07.29.26359262

**Authors:** John W. Ayers, Adam Poliak, Alexandra DeLucia, Zechariah Zhu, Stephanie R. Pitts, Mario A. Navarro, Sharareh Shojaie, Mark Dredze

**Author notes:** **Corresponding Author:** John W. Ayers, Ph.D. M.A.; #333 CRSF 9500 Gilman Drive, La Jolla, California 92093; Website: www.johnwayers.com.

## Abstract

While the public perceives e-cigarettes as less harmful than combustible tobacco, little is known about their specific health concerns regarding vaping. We demonstrate a data-driven strategy to discover the public’s health concerns about vaping e-cigarettes expressed on social media. We obtained all public posts from the largest e-cigarette-related subreddit, r/electronic_cigarette, from its inception on September 17, 2008, through April 1, 2022 (N = 10,403,433). We identified health concerns attributed to vaping by (a) selecting all cause phrases containing “cause” and its inflections, (b) calculating the empirical frequency ratio of words and bi-grams occurring in these phrases relative to random phrases, (c) retaining the 10% of words with the greatest empirical frequency of occurring in cause phrases, and (d) annotating this sample for health-relevant concerns and their subjects. In total, 76,342 posts contained cause phrases, with increased volume over time. Of the 425 words most strongly associated with cause phrases compared to random phrases, 53.4% (95%CI, 48.7-58.1) were identified as health-relevant. The top health-related concern was lipoid pneumonia, cited in 5.9% (95%CI, 5.0-6.8) of all cause phrases, followed by pneumonia (4.1%; 95%CI, 3.3-4.9), and nausea (2.7%;95%CI, 2.0-3.4). The top health concern subjects were respiratory, representing 23.7% (95%CI, 18.5-29.5) of all cause phrases, followed by gastrointestinal (12.7%; 95%CI, 8.8-17.2) and cardiovascular (8.5%; 95%CI, 5.3-12.3) concerns. Other subjects included neurological, dermatological, oral health, sexual health, psychiatric, oncologic, addiction, and sleep concerns. Because our strategy relies on data-driven techniques, our analysis can be integrated into routine social media monitoring and applied across different types of social media and text data, potentially leading to more timely identification of emerging concerns and a broader understanding across platforms. As a result, experts can craft messaging that accounts for current perceptions held by the public using our method.

## Introduction

Public health campaigns aimed at discouraging tobacco use frequently focus on reinforcing users’ understanding of and highlighting previously unknown risks of tobacco use (1,2). For instance, the inaugural Tips from Former Smokers Campaign helped educate people who smoke about health concerns, including some that might have been previously unknown to them. This increased knowledge about tobacco-related health risks was linked to higher smoking cessation rates (3,4). Similarly, understanding the public’s vaping-related health concerns is crucial to public health efforts, as it helps experts craft messaging that aligns with current public perceptions (5).

Existing methods for uncovering the public’s health concerns (6,7) – including surveys, interviews, and focus groups – provide valuable data but can face inherent challenges (8). First, they prime respondents with questions that elicit social desirability bias, restrict response options, or raise concepts that are not otherwise salient to the respondent. Second, rising costs and concerns about overburdening respondents mean survey data can only be collected intermittently, and researchers are restricted to broad measures (e.g., asking whether vaping is riskier than smoking with a simple yes/no response). As a result, detailed and temporally specific patterns in health concerns remain under-appreciated.

We leverage digital data (9), such as social media posts, to overcome these limitations and provide naturalistic and real-time insights into the public’s health concerns regarding vaping (10–12). However, applying traditional manual methods, such as intensive qualitative analyses, to digital data can be time-consuming and limited in scope. For instance, researchers can spend hundreds of hours simply annotating data for a single analysis using qualitative techniques, such as thematic coding or grounded theory analysis (13). To make digital data analyses more feasible, we demonstrate a data-driven strategy to extract the health concerns that the lay public attributes to vaping nicotine from social media posts, facilitating more timely identification of emerging concerns and broader understanding across platforms.

## Methods

### Ethics Statement

This study was conducted using publicly available and publicly forward-facing data. As such, it does not involve human participants or identifiable private information and was deemed exempt from ethical review by the Johns Hopkins University Institutional Review Board (45 CFR §46).

Reddit, a social media platform boasting 430 million monthly active users, features topically focused forums known as subreddits (14). We chose Reddit for our study for three reasons: 1) topical subreddits provide specificity; 2) Reddit posts are longer than posts on other social media platforms, offering a richer context for analysis, and; 3) Reddit has a relatively low prevalence of advertising spam compared to other social media platforms. Our research centered on the largest public e-cigarette-focused subreddit: r/electronic_cigarette. This ensured that our findings would primarily pertain to vaping nicotine rather than other substances. Using the Pushshift API (github.com/pushshift/api), we gathered all 10,403,433 posts—including original submissions and replies—from the subreddit’s inception on September 17, 2008 through April 1, 2022.

We isolated posts potentially attributing vaping as a cause for health concerns. First, we identified all posts containing “cause” and alternative inflections of the verb, such as “caused,” “causing,” or “causes.” We used “cause” phrases because they have a high degree of precision for describing cause claims. Next, we extracted the cause phrases, which included each instance of the word “cause” (or its inflection) and up to five subsequent words. Examples included “cause asthma?”, “causes cancer,” or “cause me to cough,” among other cause phrases. As a comparator, we generated a random set of 100,000 five-word phrases from posts that did not include “cause” or its inflections, using the same method.

We adopted a data-driven approach to pinpoint words significantly more likely to appear in cause phrases compared to random phrases. Initially, we utilized the Natural Language Toolkit WordNet Lemmatizer (15) to condense the vocabulary or total unique words in the posts by lemmatizing each word. Lemmatizing involves clustering words into their root form, such as converting “coughing,” “coughs,” and “coughed” into “cough.” Subsequently, we calculated the empirical frequency ratio for a lemmatized word to appear in a cause phrase compared to a random phrase, based on the rates of each word appearing in either type of phrase. A word with an empirical frequency ratio of 10, for example, is 10 times more likely to occur in cause phrases than random phrases. Notably, some words may only appear in cause phrases and never in random phrases, resulting in an infinite empirical frequency ratio score for that word. We then ranked the lemmatized words in the vocabulary by this ratio.

Our analytical sample (the subset of data matching our inclusion criteria) was composed of words occurring in the top 10th percentile of the empirical frequency ratio score, which corresponded to a score greater than 7.64 (e.g., a word was 7.65 times more likely to occur in a cause than a random phrase). A team of investigators (A.D., A.P., Z.Z., J.W.A., M.D.) labeled these words for two concepts: (1) Was a health-relevant concern described (yes/no)? (2) Among health-relevant concerns, what was the corresponding subject category of the health concern? For the latter, our team assigned health concerns to a category using open-coding. Open-coding involves examining the data without predetermined categories and creating categories based on the data itself. Disagreements and resolution of overlapping or duplicative health categories were resolved with deliberation involving the two relevant investigators and a common third investigator (J.W.A.). When two words occurred in the same cause phrase (e.g., “lipoid pneumonia”) they were labeled as a bi-gram since they were jointly referring to a single concern. In these cases, single words comprising a bi-gram did not contribute to the results for another single word health concern (e.g., instances of “lipoid pneumonia” did not contribute to the results for “pneumonia,” but were considered distinct).

We analyzed temporal changes in cause phrase posts by dividing them into monthly and/or yearly bins for the entire period. We used an ANOVA test to assess any significant changes between time bins. Among the cause phrases we report the percent of responses where the studied word/bi-gram was health-relevant. Additionally, among the subset of health-relevant vocabulary words/bi-grams, we report the prevalence (percentage of words/bi-grams labeled as belonging to each health category). We calculated 95% confidence intervals from a bootstrap of the data. Analyses were computed using Python 3.10.0.

## Results

A total of 76,410 posts on r/electronic_cigarette contained cause phrases. The monthly rate of posts containing cause phrases per 100,000 total posts increased significantly during the study period (F=68.83, p < 0.001) (**Figure 1**). There were 669 cause phrase posts per 100,000 during 2016, 813 during 2017, 902 during 2018, 1306 during 2019, 1240 during 2020, 1531 during 2021, and 1,635 during the first 3 months of 2022 (January 1, 2022 through April 1, 2022).

**Figure 1:**
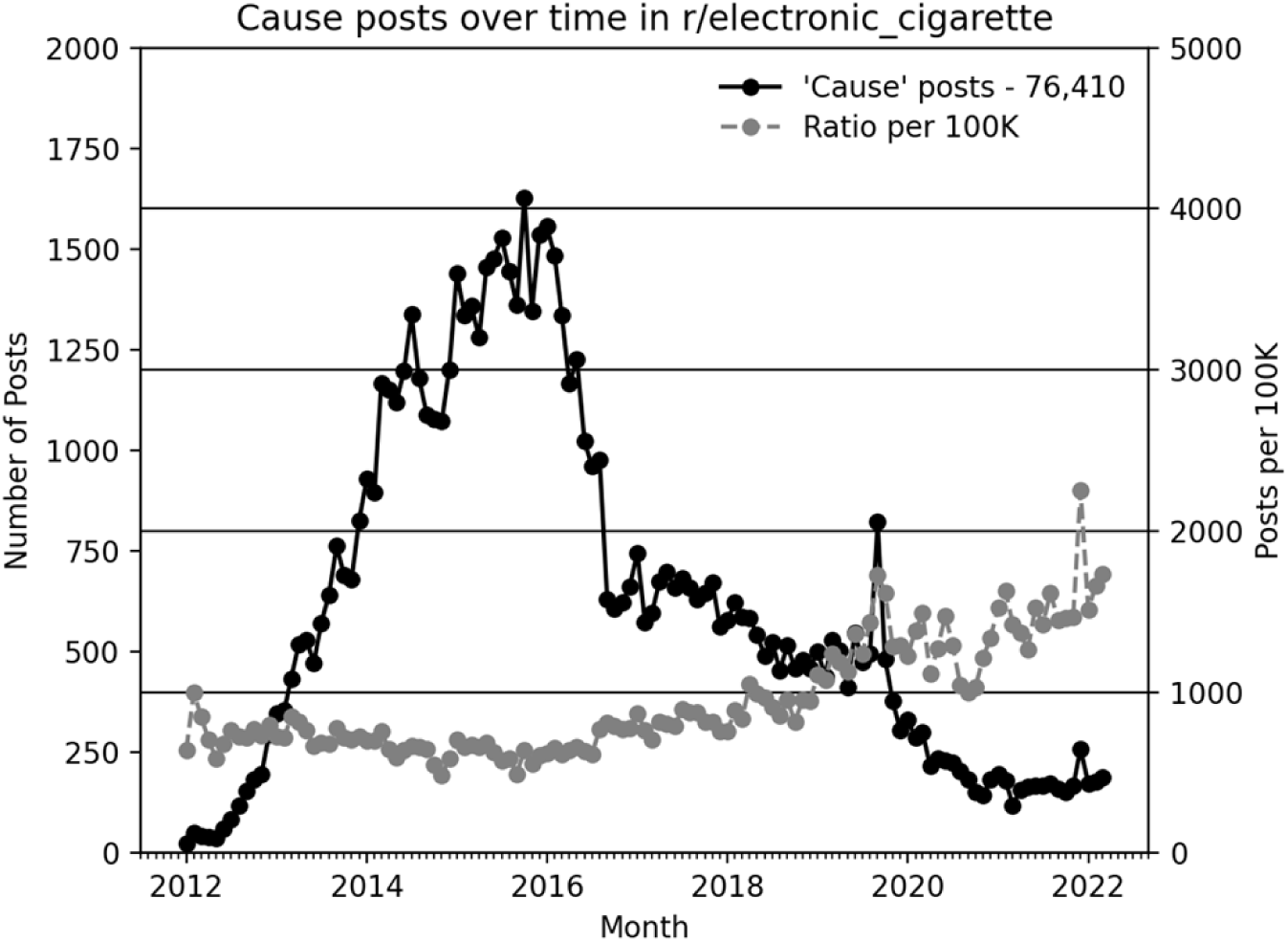
r/Electronic_cigarette posts containing cause phrases Counts are aggregated by month. The ratio is the rate of all r/electronic cigarette posts that contain cause phrases for each month per 100,000 posts. The number of posts before 2012 is negligible and near zero, and was therefore omitted from the graphic.

Among the 425 words or bi-grams within the top 10th percentile empirical frequency ratio of appearing in a cause phrase, 53.4% (95%CI, 48.7-58.1) were health-related, appearing in 2,221 cause phrases; 80.2% were single words, and the remaining 19.8% were bi-grams discovered when two words occurred repeatedly in the same cause phrase. The most frequently mentioned health-related concern in cause phrases was “lipoid pneumonia”, cited in 5.9% (95%CI, 5.0-6.8) of all health-relevant cause phrases, followed by “pneumonia” (4.1%; 95%CI, 3.3-4.9) and “nausea” (2.7%; 95%CI, 2.0-3.4) (**Figure 2**). Additional specific concerns included words relevant to a broad array of health concerns, ranging from “cavity” to “rash.” Open-ended clustering of the specific concerns yielded 11 categories of health concerns. Respiratory concerns were the most cited category of health-related concerns, encompassing 23.7% (95%CI, 18.5-29.5) of all health-related cause phrases, including “obliterans,” “bronchitis,” “emphysema” among other words and their inflections (**Table 1**). Gastrointestinal (12.7%; 95%CI, 8.8-17.2) and cardiovascular (8.5%; 95%CI, 5.3-12.3) concerns followed, being cited nearly half as much as respiratory concerns, and included concerns such as “heartburn” and “palpitation.” Neurological (6.0%; 95%CI, 3.1-8.8), dermatological (5.6%; 95%CI, 2.6-9.3), oral health (4.4%; 95%CI, 2.2-7.5), and sexual health (4.3%; 95%CI, 1.8-7.0) represent the third most common categories of concern. Psychiatric concerns were cited in 2.6% (95%CI, 0.9-4.8) of cause phrases. Oncology, addiction, and sleep concerns were each cited in less than 2% of all health-related cause phrases.

**Figure 2:**
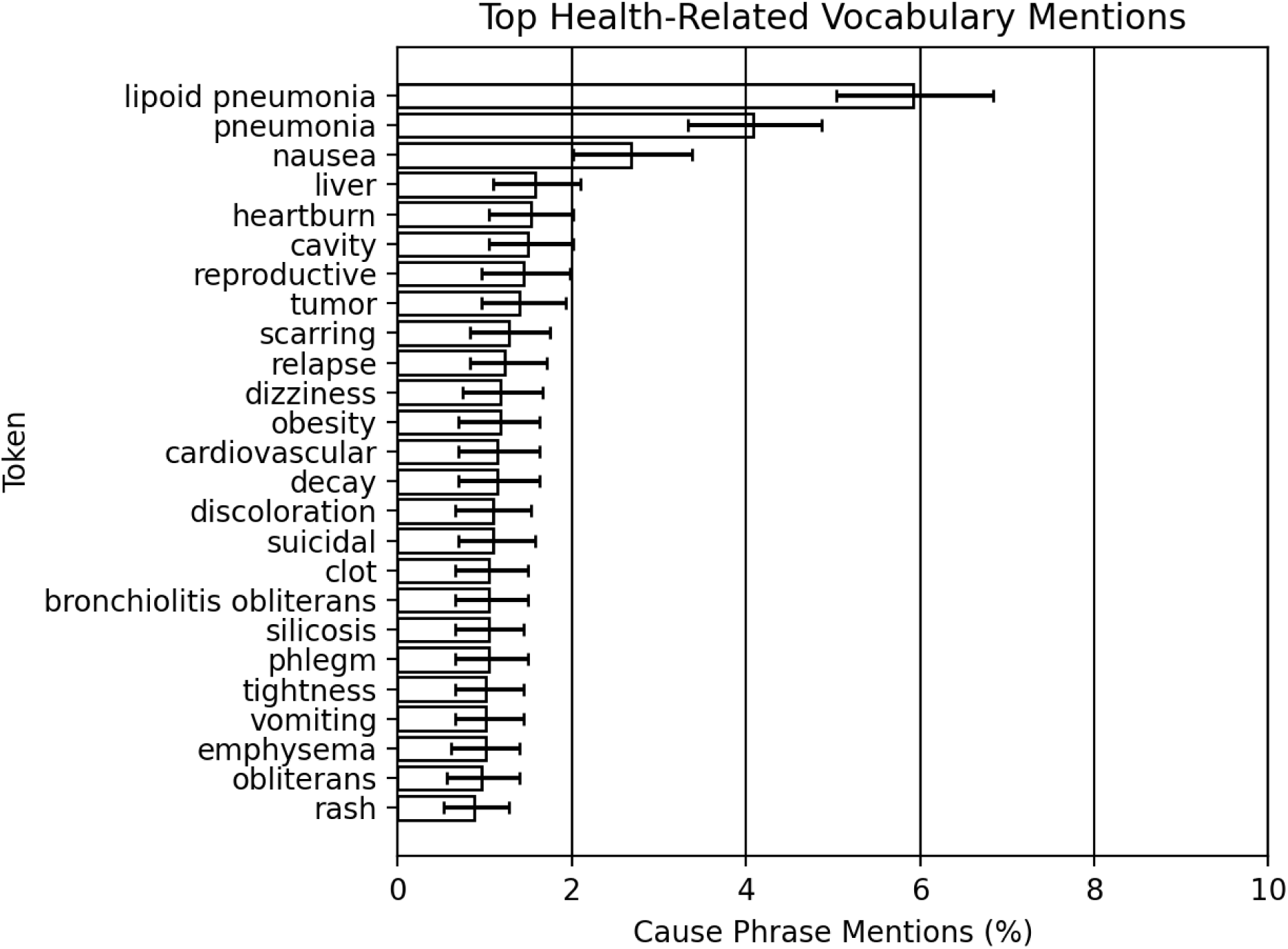
Specific health concerns raised in cause phrases on r/Electronic_cigarette. The figure presents the most frequently mentioned health-related words and bi-grams in cause phrases, shown as a percentage of their occurrence relative to all health-relevant cause phrases in the analytic sample.

**Table 1.** Categories of Health-Related Cause Concerns Reported on Reddit r/electronic_cigarettes.

| Category | Example Cause Phrases* | Prevalence (95%CI)** |
| --- | --- | --- |
| Respiratory | Aren't we realizing now that ANY oil vaped into the lungs is a horribly bad idea? And can **cause that lipoid pneumonia or some** other parade of medical disaster? | 23.7% (95%CI, 18.5-29.5) |
| Gastrointestinal | The same question has been asked, but no one has ever come back to update if they ever got a definitive answer. Can vaping **cause acid reflux**? Does smoking cigarettes cause acid reflux? The two involve inhalation and nicotine. The nicotine gets into your blood. Acid reflux starts in the stomach. There does not appear to be a correlation. | 12.7% (95%CI, 8.8-17.2) |
| Cardiovascular | When a dry hit occurs, vape pens release incredibly toxic and powerful proven carcinogens known as aldehydes, including formaldehyde. It could potentially cost you your life if you aren't careful and have a heart problem. Link to the pubmed article proving release of aldehydes here:<br>http://www.ncbi.nlm.nih.gov/pubmed/25996087 Link to article showing acetaldehyde as **cause for cardiac arrhythmia**<br>http://www.ncbi.nlm.nih.gov/pubmed/2883944 | 8.5% (95%CI, 5.3-12.3) |
| Neurological | People don't talk enough about the dangers of ecigs. Using the high nicotine levels, even as low as 12mg can allow you, if you chain vape, to overdose on nicotine enough to **cause permanent tinnitus**. At least one woman has been found with a fatal level of nicotine in her system from using these.<br>Very dismissive response, but I hope you've tried drinking significantly more water while vaping and seeing if it makes a difference. This is more than likely the cause of the dizziness at least.<br>Sounds like you might actually have a bug. Normally only increasing nicotine consumption will cause dizziness and headaches.<br>10mg is high depending on the setup for someone that's never used nicotine before. It's too much for | 6.0% (95%CI, 3.1-8.8) |
|  | you and can also cause headaches, dizziness, and nausea. If you're dead set on using nicotine, try 3mg, I'd advise against it though<br>"Only about 1 in 1,000 people are sensitive to vegetable glycerin (VG). Allergic reactions to VG can cause itching, shortness of breath, dizziness, nausea and diarrhea however, it is important to note that some of these symptoms can also be caused by tobacco withdrawal, a condition sometimes called "smoker's flu" because the symptoms resemble influenza. Talk to your doctor if you are concerned about a VG allergy." |  |
| Dermatological | potential side effect people actually have an allergy to will normally **cause itchiness rash and possibly hives** often claim they get headache etc with not so sure about that people also react badly with certain chemical in some flavoring such a cinnamon and creamy flavoring Ive been looking into this bit myself a pretty sure have allergy but don want to discount anything else | 5.6%<br>(95% CI, 2.6-9.3) |
| Oral Health | Yup, sweetened vapes are more likely to **cause cavities or calculus hardening**. Though it confused me as they said alcohol ethers are more likely to cause the breakdown process, but that sugar alcohols inhibit the growth of a bacteria. Also happy cake day :o | 4.4%<br>(95%CI, 2.2-7.5) |
| Sexual Health | Sometimes having a place to lay the blame, even if it is on your own shoulders, is better than not having any place to put it down. Nicotine did not **cause the miscarriage**. You did not cause it. It just happens. | 4.3%<br>(95%CI, 1.8-7.0) |
| Psychiatric | On top of that, the nicotine hit itself, if you are a nicotine addict as most of us are, will be interpreted by the brain as a "pleasurable experience" and **cause dopamine release which will fire** up the reward circuitry of the brain. So nicotine enhances the vaping experience and can lead you to perceive any flavor as more enjoyable. | 2.6%<br>(95%CI, 0.9-4.8) |
| Oncologic | You put in zero, because you don't smoke. Nicotine is not a recognized as a carcinogen, even though there have been many studies on it. It appears to be what they call a tumor promoter, but it does not appear to **cause tumors itself**. | 1.8%<br>(95%CI, 0.4-3.5) |
| Addiction | Back to vaping on El Kamino, or Citrus Cooler, or | 1.5% |
|  | any of the other 50 flavors I keep around. **Cause I am a junkie for** flavors. | (95%CI, 0.4-3.1) |
| Sleep | I've read studies online that say high dose nicotine can **cause insomnia**. But they are talking like 500+ mg I believe? | 0.4%<br>(95%CI, 0.0-1.3) |
| * Example phrases are shown in bold text with the surrounding content available for interpretation.<br>** Prevalence refers to the proportion of all health-related cause phrases belonging to the category. |  |  |

## Discussion

Our analysis revealed a range of health concerns that users associate with vaping and share publicly. The most frequently mentioned specific health concerns were lipoid pneumonia, pneumonia, and nausea. The top categories of health concerns were respiratory concerns, followed by gastrointestinal and cardiovascular concerns, with eight additional categories of concerns.

Our data-driven method minimizes preconceived biases and assumptions, allowing for the discovery of unanticipated health concerns (16). This can augment traditional methods and promote scalability and reproducibility when incorporated into ongoing public health surveillance alongside other data sources. The latter can be particularly useful in dynamic arenas such as vaping, which undergoes rapid changes in use rates, communication, or regulation. The strengths of our method complement traditional survey approaches providing a more comprehensive understanding of vaping health concerns when combined with other surveillance data. Last, the strategy herein is scalable to other types of tobacco products, social media, and text data.

By analyzing social media, we also discovered areas of self-reported concern that may have been overlooked in research using traditional forms of data such as survey data. This includes concerns about gastrointestinal or psychiatric concerns that Reddit users related to vaping (17,18). Self-reported narratives can provide a unique and unmediated perspective on health concerns that might influence their vaping behaviors. Exploring these concerns can offer context on beliefs and perceptions to inform the design of complementary research and evidence-based strategies to educate the public about vaping. It also can allow researchers to identify gaps in knowledge or misperceptions about product effects.

Real-time implementation of our strategy can help public health professionals stay ahead of emerging issues and respond to public concerns in a timely manner, such as with e-cigarette, or vaping, product use-associated lung injury (20–22). Similarly, just as social media is a venue for understanding the public, it also is a venue for educational interventions. Responses to cause-related posts could be tested for their efficacy as a pilot for larger communication campaigns. For instance, cause phrases could be responded to using chatbots (such as adaptations of ChatGPT (23)) and the resulting interaction with the original poster and other commenters could inform the evaluation of communication strategies that most effectively engage and educate tobacco users (24).

Our study used Reddit, one of the most popular social media platforms. Reddit’s user demographics do not reflect the general U.S. population, as the platform skews younger and more male-dominated than other social media sites. However, this demographic distribution closely aligns with common users of e-cigarettes, who are primarily young adults ages 18-24. Studies show that this age group has the highest prevalence of e-cigarette use, with many initiating vaping without prior use of combustible cigarettes, making Reddit a particularly relevant platform for examining discussions related to vaping (26). Additionally, the comments in posts reflect users who are active online in discussing these topics, which may differ from those of less vocal individuals.

The study was limited to a single subreddit focused on e-cigarettes; we did not verify that all posts studied were specifically about vaping nicotine. We only observed “cause” phrase concerns. Concerns described using other keywords may differ from the results reported herein. Additionally, a portion of cause phrases may not be health-related and necessitates the curation of our results by human annotators. Concerns raised in online forums may not be supported by scientific evidence and may not reflect the actual health risks associated with vaping. However, our focus was on understanding perceptions (25). Public health professionals work to educate the public, which requires them to understand existing perceptions of these products. Moreover, we studied social media posts in aggregate and did not link potential health concerns with any demographic group or geolocation. Last, the population of Reddit users may not be representative of e-cigarette users in the United States as a whole. However, the study has several strengths, notably those related to our data-driven method compared to a traditional hypothesis-driven analysis.

In the future, our method could be implemented for near real-time health concern detection with expansion across different types of social media and potential tobacco products, potentially becoming a supplemental source for routine tobacco control intelligence.

## Acknowledgement

This work was supported by the U.S. Food and Drug Administration (FDA) via the Johns Hopkins University Center of Excellence in Regulatory Science and Innovation (CERSI Grant #UFD005942B, FOA: RFA-FD-18-011; J.W.A., A.P., A.D., Z.Z., and M.D.). Additional funding was provided by the Burroughs Wellcome Fund (1017617.01; J.W.A., Z.Z., and M.D.). FDA (S.R.P., M.A.N., and S.S.) played a central role in study design, data collection and analysis, decision to publish, and preparation of the manuscript. The Burroughs Wellcome Fund had no role in study design, data collection and analysis, decision to publish, or preparation of the manuscript. The findings and conclusions in this report are those of the authors and do not necessarily represent the official position of FDA. M.A.N. worked on this manuscript while employed at the U.S. Food and Drug Administration’s Center for Tobacco Products. M.D. had full access to all the data in the study and takes responsibility for the integrity of the data and the accuracy of the data analysis.

## Data Availability Statement

The data used in the study are public in nature. Reddit prohibits investigators from providing raw data to anyone outside of the research team.

## Author Contributions

JWA, AD, AP, SRP, MAN, SS, MD led the study design. AP, AD, and MD led data collection and JWA, AD, AP, ZZ, and MD led the data analysis. All authors participated in the drafting of the manuscript and agreed to the final submission.

## References

1. U.S. Food and Drug Administration. Think E-Cigs Can’t Harm Teens’ Health? April 30, 2020. Accessed May 9, 2023. https://www.fda.gov/tobacco-products/ctp-newsroom/fdas-comprehensive-plan-tobacco-and-nicotine-regulation.

2. U.S. Food and Drug Administration. Vaporizers, E-Cigarettes, and Other Electronic Nicotine Delivery Systems (ENDS). September 17, 2020. Accessed May 9, 2023. https://www.fda.gov/tobacco-products/products-ingredients-components/vaporizers-e-cigarettes-and-other-electronic-nicotine-delivery-systems-ends

3. Ayers JW, Althouse BM, Emery S. Changes in Internet searches associated with the “Tips from Former Smokers” campaign. Am J Prev Med. 2015;48(6):e27–e29.

4. Davis KC, Duke J, Shafer P, Patel D, Rodes R, Beistle D. Perceived Effectiveness of Antismoking Ads and Association with Quit Attempts Among Smokers: Evidence from the Tips From Former Smokers Campaign. Health Communication. 2017;32(8):931–938.

5. Bailey LS, Stewart S, Hatala J, et al. Mass media interventions for smoking cessation in adults. Cochrane Database Syst Rev. 2014;11(11):CD004704.

6. Ma H, Gaudiello E, Sheeran P, Sanzo N, Sutfin EL, Noar SM. National youth tobacco surveys (2014-2019) show increasing beliefs in the harm and relative addiction of e-cigarettes but decreasing associations between those beliefs and e-cigarette use. Addict Behav. 2023;144:107713

7. Grummon AH, Hall MG, Mitchell CG, Pulido M, Sheldon JM, Noar SM, Ribisl KM, Brewer NT. Reactions to messages about smoking, vaping and COVID-19: two national experiments. Tob Control. 2022;31(3):402–410.

8. Zaller J, Feldman S. A simple theory of the survey response: Answering questions versus revealing preferences. Am J Pol Sci. 1992;36(3):579–616.

9. Ayers JW, Althouse BM, Dredze M. Could behavioral medicine lead the web data revolution? JAMA. 2014;311(14):1399–1400.

10. Ayers JW, Althouse BM, Ribisl KM, Emery S. Digital detection for tobacco control: Online reactions to the United States’ 2009 cigarette excise tax increase. Nicotine Tob Res. 2013;16:576–583.

11. Ayers JW, Leas EC, Allem JP, et al. Why do people use electronic nicotine delivery systems (electronic cigarettes)? A content analysis of Twitter, 2012-2015. PloS One. 2017;12:e0170702.

12. Ayers JW, Dredze M, Leas EC, Caputi TL, Allem JP, Cohen JE. Next generation media monitoring: Global coverage of electronic nicotine delivery systems (electronic cigarettes) on Bing, Google and Twitter, 2013-2018. PloS one 2018;13(11):e0205822.

13. Paul MJ, Dredze M. Social monitoring for public health. Synthesis Lectures On Information Concepts, Retrieval, and Services. Vol. 9, pp. 1–183. Morgan & Claypool Publishers; 2017 .

14. Curry D. Reddit revenue and usage statistics (2022). Business of Apps. Published October 2, 2020. Accessed August 29, 2022. https://www.businessofapps.com/data/reddit-statistics/

15. Bird S, Loper E, Klein E. Natural Language Processing with Python. 2009; O’Reilly Media Inc.

16. Brunton SL, Kutz JN. Data-Driven Science and Engineering. Cambridge, UK: Cambridge University Press; 2019.

17. Noar, SM, Jang Y, Zarndt AN, Zhao X, Ross JC, Cappella JN. Achieving public health impact: Health communication research to inform tobacco regulatory science. Health Comm. 2024;10.1080/10410236.2024.2326250

18. Ayers JW, Leas EC, Dredze M, Caputi TL, Zhu SH, Cohen JE. Philip Morris International used the e-cigarette, or vaping, product use associated lung injury (EVALI) outbreak to market IQOS heated tobacco. Tob Control. 2021;doi:10.1136/tobaccocontrol-2021-056661.

19. U.S. Food and Drug Administration. The Real Cost Campaign. February 26, 2021. Accessed May 9, 2023. https://www.fda.gov/tobacco-products/public-health-education/real-cost-campaign.

20. King BA, Jones CM, Baldwin GT, Briss PA. The EVALI and youth vaping epidemics— implications for public health. N Engl J Med. 2020;382(8):689–691.

21. Leas EC, Nobles AL, Caputi TL, et al. News coverage of the e-cigarette, or vaping, product use associated lung injury (EVALI) outbreak and Internet searches for vaping cessation. Tob Control 2021;30:578–82.

22. Tattan-Birch H, Brown J, Shahab L, Jackson SE. Association of the US outbreak of vaping-associated lung injury with perceived harm of e-cigarettes compared with cigarettes. JAMA Netw Open. 2020;3(6):e206981.

23. Ayers JW, Zhu Z, Poliak A, et al. Evaluating artificial intelligence (ChatGPT) responses to public health questions. JAMA Network Open. Published online June 5, 2023. doi:10.1001/jamainternmed.2023.1838

24. Whittaker R, Dobson R, Garner K. Chatbots for smoking cessation: scoping review. J Med Internet Res 2022;24(9):e35556.

25. National Academies of Sciences Engineering and Medicine. Public Health Consequences of E-Cigarettes. Washington, DC: National Academies Press; 2018.

26. Erhabor J, Boakye E, Obisesan O, et al. E-cigarette use among US adults in the 2021 Behavioral Risk Factor Surveillance System Survey. JAMA Netw Open. 2023;6(11):e2340859. doi:10.1001/jamanetworkopen.2023.40859

